# Neuroimaging Insights into Subjective Cognitive Decline: Differential Sensitivity of Cognitive Change Index and Everyday Cognition Scale

**DOI:** 10.1101/2024.06.10.24308700

**Authors:** Cassandra Morrison, John A. E. Anderson, Mahsa Dadar, the Alzheimer’s Disease Neuroimaging Initiative

## Abstract

**Introduction:** Cognitively healthy older adults may experience self-perceived memory and cognitive deficits, known as subjective cognitive decline (SCD), increasing their risk for dementia-related brain and cognitive changes. This study investigated if questions from the Cognitive Change Index (CCI) and Everyday Cognition Scale (ECog) show similar associations with dementia-related changes.

**Methods:** Cognitively healthy older adults (n=332) from the Alzheimer’s Disease Neuroimaging Initiative were included. Partial-least-squares observed the latent variables (LVs) that maximize the relationship between the two questionnaires.

**Results:** Two LVs (*p’s*<0.001) explained 85.89% and 8.30% of the cross-block covariance. In the first LV, several CCI questions correlated with older age and frontal, parietal, and temporal WMHs, lower hippocampal and entorhinal cortex volume, and larger ventricles. The second LV showed younger individuals with higher SCD scores on three CCI questions correlated with temporal and parietal WMHs and entorhinal cortex volumes.

**Conclusion:** More questions from the CCI are associated with neuroimaging markers, unlike the ECog questions. These questionnaires may thus be measuring different neural decline patterns and may be sensitive to different etiologies.

## 1. Introduction

A common phenomenon observed in cognitively healthy older adults is subjective cognitive decline (SCD) or self-reported declines in memory or cognitive functioning without objective changes on standardized cognitive tests (Rabin et al., 2017). Cognitively healthy older adults who experience SCD exhibit increased rates of cognitive decline(Morrison and Oliver, 2022), neurodegeneration (Jessen et al., 2006; Striepens et al., 2010), white matter hyperintensity (WMH) burden (Morrison et al., 2023; Rooden et al., 2018), and Alzheimer’s disease (AD)-related pathology (Perrotin et al., 2017; Snitz et al., 2015), compared to cognitively healthy older adults who do not experience SCD (for review see Wang et al., 2020). These group differences have led to the understanding that SCD may be the *preclinical* stage of AD (Jessen et al., 2020). Therefore, much research has investigated SCD to improve early AD detection (Rabin et al., 2017; Wang et al., 2020), which would allow for interventions and treatment options to be successful before substantial pathological brain changes have occurred (Gauthier et al., 2016; Sperling and Aisen, 2011).

Despite this vast body of research on SCD, there is no consistent definition of how researchers (or clinicians) classify people with SCD. That is, different strategies for SCD assessment ranging from a single question such as, “Do you feel your memory is becoming worse?” to a more comprehensive questionnaire assessment including domains such as memory, language, executive functioning, and visuospatial abilities have been employed. Standard validated tests to gather comprehensive data on cognitive complaints include the Cognitive Change Index (CCI, Saykin et al., 2006), Everyday Cognitive Scale (ECog, Farias et al., 2008), and the Memory Assessment Clinics Questionnaire (MAC-Q, Crook et al., 1992). In addition to these questionnaires, endorsement of “Worry” about one’s cognitive change may be an important factor to consider when examining these assessment techniques. Other factors that increase the risk of AD include genetic risk for AD (APOE e4 positivity), SCD onset after 60 years of age and within the last five years, and feelings of worse performance compared to others (Jessen et al., 2020).

Variability in the methods used to identify SCD could have serious implications for current findings examining the relationship between SCD and brain and cognitive changes. For example, one study examining the relationship between the Subjective Memory Complaints Scale and The Memory Complaint Questionnaire only found a moderate correlation (R = 0.44, Vogel et al., 2016), while another study comparing the Blessed memory test to the full ECog and to the memory subset of the ECOG found slightly higher correlations (R=0.52 and R=0.50, van Harten et al., 2018), respectively. These correlations between the questionnaires suggest that they are not interchangeable and may measure different underlying constructs. In previous findings, we examined atrophy, WMHs, and cognition in cognitively healthy older adults with and without SCD using four different SCD assessment methods (including the CCI, ECog, ECog+Worry, and Worry alone). We observed that the four methods used to classify SCD resulted in distinct brain and cognitive change patterns. For example, WMH burden was higher in temporal and parietal regions in people with SCD using both the CCI and ECog (Morrison et al., 2023). However, while SCD defined using the CCI was associated with atrophy in the left hippocampus, the ECog was associated with atrophy in the left and right superior temporal regions (Morrison et al., 2022). These findings suggest the different methods used to assess SCD may reflect different underlying pathologies and subsequent cognitive decline. A recent study by Rabin and colleagues (2023) harmonized self-report questionnaire data from over 20 studies and 40 different questionnaires using item-response theory. They observed that a single-factor structure was particularly valuable for assessing different aspects of cognitive functioning and was reasonable for the latent trait of SCD. However, because cognitive symptoms of AD occur up to 10-20 years after brain changes (Jessen et al., 2020; Reisberg et al., 2010), it is critical to examine whether questionnaires equally contribute to structural brain changes that may signal future cognitive decline.

Two other major factors that have shown important relationships with SCD but are underexplored are biological sex and cognitive reserve. For example, previous findings indicate that SCD is more predictive of cognitive decline in females than males (Morrison and Oliver, 2022) and that WMHs influence cognitive decline more in females than males, even when they exhibit the same amount of WMH loads (Morrison et al., 2023). We thus wanted to determine if participant sex alters the relationship between SCD and structural brain changes associated with these questionnaires. Furthermore, SCD has also been shown to be influenced by cognitive reserve (CR). Cognitive reserve refers to an individual’s ability to defy age or disease-related neuropathology and cognitively outperform their apparent brain state – likely via increased functional and structural connectivity, which allows them to circumvent local damage (Stern, 2013, 2002). Cognitive reserve has been associated with lifetime measures of socio-behavioural proxies such as higher education, musicianship, and speaking multiple languages and can extend the period of cognitive normalcy before diagnosis by up to five years (Anderson et al., 2020; Stern, 2002; Wilson et al., 2015). People with high CR thus cope better with similar amounts of pathology than individuals with low cognitive reserve. Higher levels of educational attainment have also been shown to be associated with increased rates of amyloid burden and conversion to AD in people with SCD (Aghjayan et al., 2017). While seemingly paradoxical, a faster level of decline in individuals with higher CR is consistent with a portrait of a brain that has staved off cognitive decline through increasingly intricate connections between regions before neuropathological damage becomes insurmountable and the individual suffers total system collapse. Contrast this with an individual with low CR whose cognitive status declines in lock-step with incremental damage. The latter individual declines slowly but has a far longer period of cognitive dysfunction.

The current paper investigates two commonly used methods to assess SCD, the CCI and the ECog. We have previously observed different patterns of gray matter atrophy and WMH burden in people who endorse these questionnaires (Morrison et al., 2023; Morrison et al., 2022). We wanted to further examine if the individual questions that examined memory performance from these two questionnaires were associated with brain volume and WMHs to the same degree. That is, is one of the questionnaires or certain questions from the questionnaires more sensitive to brain changes indicative of cognitive decline and dementia. We also wanted to investigate whether biological sex and CR (measured through educational attainment) alter the relationship between SCD and structural brain changes in these questionnaires.

## 2. Methods

### 2.1 Alzheimer’s Disease Neuroimaging Initiative

Data used in the preparation of this article were obtained from the Alzheimer’s Disease Neuroimaging Initiative (ADNI) database (adni.loni.usc.edu). The ADNI was launched in 2003 as a public–private partnership, led by Principal Investigator Michael W. Weiner, MD. The primary goal of ADNI has been to test whether serial magnetic resonance imaging (MRI), positron emission tomography (PET), other biological markers, and clinical and neuropsychological assessment can be combined to measure the progression of mild cognitive impairment (MCI) and early Alzheimer’s disease (AD). Participants were between 55 and 90 years old at the time of recruitment. The study received ethical approval from the review boards of all participating institutions. Written informed consent was obtained from participants or their study partner. Participants were selected from the ADNI-2 and ADNI-3 cohort because ADNI2 was the first cohort to introduce the CCI. In ADNI the CCI questionnaire was used to define participants with significant memory concerns. For consistency with current research standards, we use the term subjective cognitive decline.

### 2.2 Participants

ADNI participants were between 55-90 years of age and were included if they were cognitively normal at baseline. That is, they exhibited no evidence of memory decline, as measured by the Wechsler Memory Scale and no evidence of impaired global cognition as measured by the Mini Mental Status Examination (MMSE) or Clinical Dementia Rating (CDR). Participants were included if they completed both the CCI and ECog questionnaire, had an MRI completed within 6 months from the time of the questionnaires, from which WMHs, ventricle, hippocampal, and entorhinal cortex volumes could be extracted or had at least one of the cognitive tests available including the Montreal Cognitive Assessment (MOCA), Mini Mental Status Examination (MMSE), Alzheimer’s Disease Assessment Scale-13 (ADAS-13), or Rey Auditory Verbal Learning Test (RAVLT). A total of 334 cognitively healthy older adults (212 females, 122 males) in ADNI-2 and ADNI-3 who met the inclusion criteria were included in this study.

### 2.3 Subjective Cognitive Decline Questionnaires

The questions from the two questionnaires related to memory were extracted and examined. These questions are presented in Table 1. The ECog had 8 questions and the CCI had 12 questions related to memory functioning.

**Table 1.** Items from the SCD questionnaires. Note that the “Item” label corresponds to the labels used in the PLS analyses.

| Questionnaire | Item | Question |
| --- | --- | --- |
| ECog | PT MEMORY1 | Remembering a few shopping items without a list |
| ECog | PT MEMORY2 | Remembering things that happened recently (such as recent outings, events in the news) |
| ECog | PT MEMORY3 | Recalling conversations a few days later |
| ECog | PT MEMORY4 | Remembering where he/she has placed objects |
| ECog | PT MEMORY5 | Repeating stories and/or questions |
| ECog | PT MEMORY6 | Remembering the current date or day of the week |
| ECog | PT MEMORY7 | Remembering he/she has already told someone something |
| ECog | PT MEMORY8 | Remembering appointments, meetings, or engagements |
| CCI | CCI1 | Recalling information when I really try |
| CCI | CCI2 | Remembering names and faces of new people I meet |
| CCI | CCI3 | Remembering things that have happened recently |
| CCI | CCI4 | Recalling conversations a few days later |
| CCI | CCI5 | Remembering where things are usually kept |
| CCI | CCI6 | Remembering new information told to me |
| CCI | CCI7 | Remembering where I placed familiar objects |
| CCI | CCI8 | Remembering what I intended to do |
| CCI | CCI9 | Remembering names of family members and friends |
| CCI | CCI10 | Remembering without notes and reminders |
| CCI | CCI11 | People who know me would find that my memory is |
| CCI | CCI12 | Remembering things compared to my age group |

*Notes:* ECog = Everyday Cognition Scale. CCI = Cognitive Change Index.

### 2.4 Structural MRI acquisition and processing

All scans were downloaded from the ADNI website (see http://adni.loni.usc.edu/methods/mri-tool/mri-analysis/ for the detailed MRI acquisition protocol). T1w scans for each participant were pre-processed through our standard pipeline including noise reduction (Coupe et al., 2008), intensity inhomogeneity correction(Sled et al., 1998), and intensity normalization into range [0- 100]. The pre-processed images were then linearly (9 parameters: 3 translation, 3 rotation, and 3 scaling) (Dadar et al., 2018) registered to the MNI-ICBM152-2009c average (Fonov et al., 2011).

### 2.5 WMH measurements

WMHs were generated using a segmentation technique that has been previously validated in ADNI (Dadar et al., 2019), and other multi-center studies such as the Parkinson’s Markers Initiative (Dadar et al., 2020) and the National Alzheimer’s Coordinating Center (Anor et al., 2021). Although FLAIR and T2w images are typically employed for segmentation of WMHs, we used T1w MR images because of their consistent availability across all cohorts and participants, allowing us to maximize the inclusion of the largest number of participants. Our previous work has determined that this T1w-based segmentation method holds strong correlations with the multicontrast T1w and T2w or FLAIR-based WMHs segmentations (*r* = 0.97, *p*<0.0001) and have similar relationships with clinical/cognitive scores as the multicontrast WMH segmentations (Dadar et al., 2019, 2018).

WMHs were automatically segmented using T1w contrasts along with a set of location and intensity features obtained from a library of manually segmented scans in combination with a random forest classifier to detect the WMHs in new images (Dadar et al., 2017b, 2017a). WMH load was defined as the volume of all voxels as WMH in the standard stereotaxic space (in mm3) and are thus normalized for head size. The volumes of the WMHs for frontal, parietal, temporal, and occipital lobes as well as the entire brain were calculated based on regional masks from the Hammers atlas (Dadar et al., 2017b; Hammers et al., 2003), and log-transformed to account for non-normality. The quality of the registrations and WMH segmentations was visually verified by an experienced rater (author M.D.), blinded to participants diagnostic group.

### 2.6 Freesurfer Measurements

T1w images were processed using FreeSurfer and quality controlled by the UCSF group, and regional GM volumes for the hippocampal, entorhinal cortices, and lateral ventricles were extracted. These volume estimates were collapsed across hemispheres. 1.5T and 3T data were processed with FreeSurfer versions 4.3 and 5.1, respectively, as appropriate.

### 2.7 Analysis

As multivariate analyses do not handle missing data well, before analysis, missing values were imputed using a principal components model and the missMDA R package (Josse and Husson, 2016). No variables had more than 10% of their values missing. The data were then analyzed using multivariate partial-least-squares (PLS). This method is a data-driven approach that combines elements of principal components analysis but goes further by maximizing the covariance between two data sets: one representing behavior or design (referred to as the *Y* matrix) and the other representing brain data (referred to as the *X* matrix). This analytical approach was based on the work of McIntosh & Lobaugh (2004) and Krishnan et al. (2011) (Krishnan et al., 2011; McIntosh and Lobaugh, 2004). PLS analyses were implemented in R version 4.3.1 using the Two-Table Exposition (TExPosition) package previous described in detail (Beaton et al., 2014). Thus, PLS can generate a set of orthogonal latent variables (LVs) that maximize the relationship between two data sets presented in descending order of explained cross-block covariance. Because PLS tests an entire pattern of relationships in a single step, there is no need to correct for multiple comparisons; instead, the significance of each LV was assessed by comparing the obtained result to a null distribution built with 1000 permutations. The reliability of the contributions of each of the variables to the LV was assessed with 1000 bootstrap repetitions, which were used to estimate standard errors. Salience values (i.e., the contributions for each variable) were divided by the bootstrapped standard errors to obtain bootstrap ratio (BSR) scores, which can be interpreted similarly to Z-scores. BSR scores exceeding thresholds of +/-2 were considered to reliably contribute to the LV. To examine the influence of sex and education on how SCD questions are endorsed we completed the PLS analyses again in males and females separately and then in those with high vs. low education (as determined by a median split).

We then used principal component analysis (PCA) to reduce the dimensionality across the MMSE, RAVLT forgetting, RAVLT learning, RAVLT immediate, and ADAS13 scores and produce a “memory score” from the first component. This memory component was then used as the outcome variable in two linear regressions where the predictors were the behavioral and brain scores from the PLS analysis. This analysis was completed to investigate the extent to which PLS-derived brain and SCD patterns can explain memory performance.

Receiver operating characteristic (ROC) analysis was performed to investigate the capability of the PLS LVs and PCA memory scores in predicting future cognitive decline. Cognitive decline was measured as a change of 0.5 points or greater between baseline and follow-up CDR-SB scores for one and two years following the baseline timepoint, based on which the LVs were calculated. Similar analyses were repeated with a threshold of 1 point increase in CDR-SB to assess the robustness of the results to the choice of cognitive decline threshold.

## 3 Results

Descriptive statistics by sex can be found in Table 2, which reveals that males in the sample were, on average, two years older than females and had slightly lower scores on cognitive assessments, including the MOCA, the ADAS, and RAVLT.

**Table 2.** Descriptive statistics for the healthy control sample included in the analysis.

|  | **Female**<br>**(N=212)** | **Male**<br>**(N=122)** | **Total**<br>**(N=334)** |
| --- | --- | --- | --- |
| **Age bl** |  |  |  |
| Mean (SD) | 70.3 (5.95) | 72.6 (6.16) | 71.1 (6.12) |
| Median [Min, Max] | 69.4 [55.1, 89.8] | 71.3 [61.1, 90.2] | 69.7 [55.1, 90.2] |
| **Education** |  |  |  |
| Mean (SD) | 16.7 (2.36) | 17.2 (2.19) | 16.9 (2.31) |
| Median [Min, Max] | 17.0 [8.00, 20.0] | 18.0 [12.0, 20.0] | 18.0 [8.00, 20.0] |
| **MMSE** |  |  |  |
| Mean (SD) | 29.2 (1.03) | 29.0 (1.24) | 29.1 (1.11) |
| Median [Min, Max] | 29.0 [25.0, 30.0] | 29.0 [24.0, 30.0] | 29.0 [24.0, 30.0] |
| **MOCA** |  |  |  |
| Mean (SD) | 26.4 (2.45) | 25.5 (2.60) | 26.1 (2.54) |
| Median [Min, Max] | 27.0 [19.0, 30.0] | 25.5 [19.0, 30.0] | 26.0 [19.0, 30.0] |
| **ADAS13** |  |  |  |
| Mean (SD) | 7.48 (3.86) | 10.0 (4.66) | 8.40 (4.34) |
| Median [Min, Max] | 6.67 [0.670, 21.7] | 10.0 [0, 26.3] | 7.67 [0, 26.3] |
| **RAVLT learning** |  |  |  |
| Mean (SD) | 6.64 (2.26) | 5.50 (2.60) | 6.22 (2.45) |
| Median [Min, Max] | 7.00 [2.00, 12.0] | 5.00 [-8.00, 11.0] | 6.00 [-8.00, 12.0] |
| **RAVLT immediate** |  |  |  |
| Mean (SD) | 48.6 (9.69) | 42.6 (9.50) | 46.4 (10.0) |
| Median [Min, Max] | 48.0 [25.0, 70.0] | 42.0 [25.0, 62.0] | 46.0 [25.0, 70.0] |
| **RAVLT forgetting** |  |  |  |
| Mean (SD) | 3.41 (3.70) | 4.04 (2.90) | 3.64 (3.44) |
| Median [Min, Max] | 3.00 [-24.0, 15.0] | 4.00 [-9.00, 14.0] | 3.50 [-24.0, 15.0] |

### 3.1 PLS Analysis Relating SCD to Brain Changes in Cognitively Normal Older Adults

The first PLS analysis relating SCD scores from the CCI, the ECog, and age with volumetric and white matter hyperintensity scores revealed two significant LVs, *p*’s < 0.001, which explained 85.89% and 8.30% of the cross-block covariance respectively (see Figure 1). The first LV captures a pattern where the CCI and increasing age load together along with a higher probability of WMH in all regions except occipital, larger lateral ventricle volume, and reduced gray matter volume in hippocampal and entorhinal cortices. The five questions from the CCI that loaded together with increasing age and greater neuropathology were questions 2, 6, 7, 10, and 12, which tend to have a more concrete focus (e.g., memory for faces, new information, objects, memory without notes, and a comparison to the person’s age group) than questions which did not load as strongly (e.g., recalling information, recalling conversations, and remembering what they intended to do).

**Figure 1:**
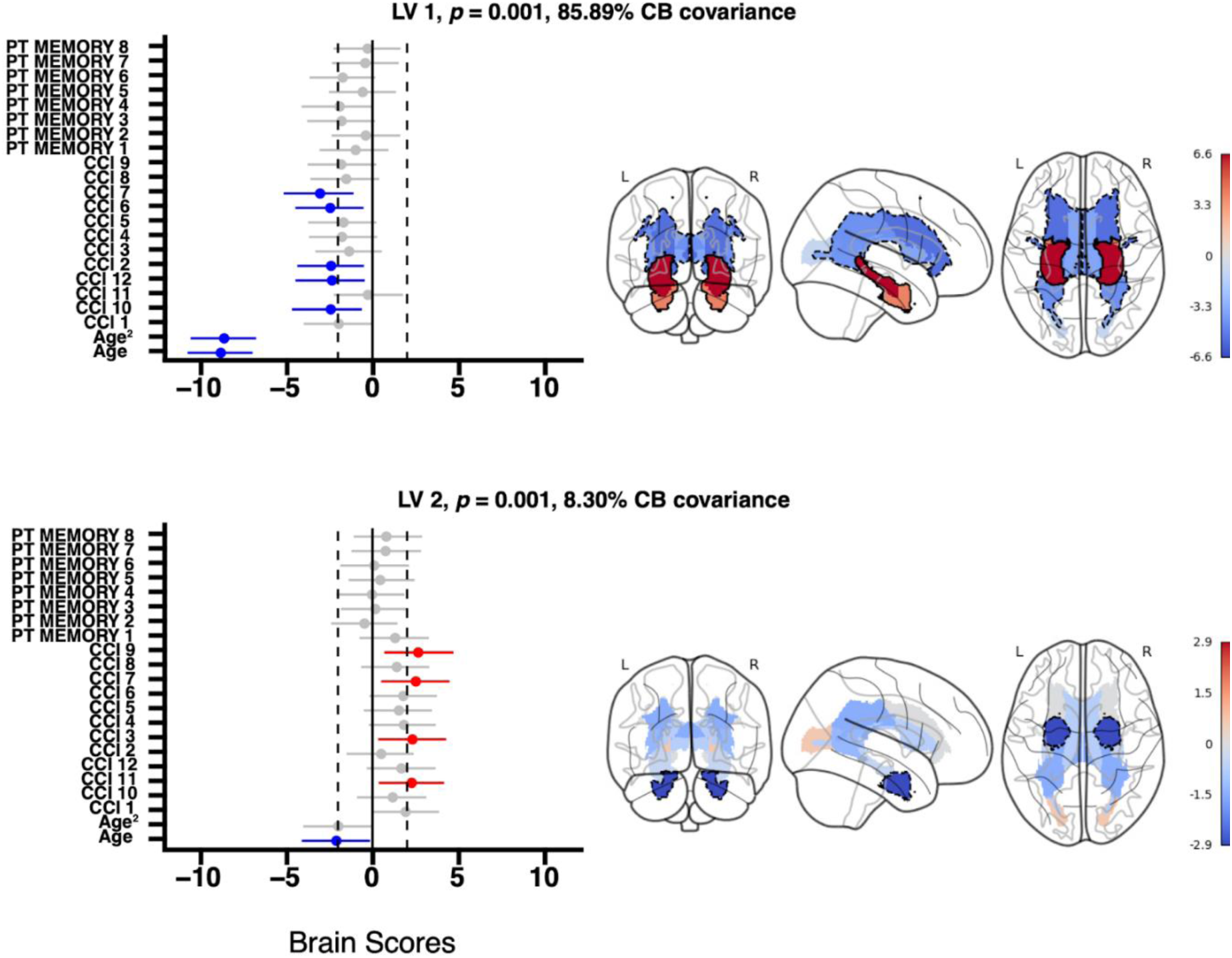
PLS analyses for full sample. *Notes:* PT_Memory 1 through 8 are the ECOG questions. CCI1 through 12 are the CCI questions. PLS Analysis results for healthy older adults from the ADNI database and links subjective cognitive decline, as measured by the Cognitive Change Index (CCI) and Everyday Cognition (ECog) scores, with volumes of various brain regions, including white matter hyperintensity, hippocampus, entorhinal cortex, and ventricles. White matter hyperintensity volumes are specifically parcellated into Hammer’s Atlas lobes, encompassing frontal, parietal, occipital, and temporal regions. The visualization includes point-range plots showing the loadings, or brain scores, for each of the questions on each latent variable. Brain images within the figure highlight bootstrap ratio values, with areas exceeding absolute values of +/-2 outlined in black for emphasis. The color coding is critical for interpretation: blue-colored regions in the brain, as well as blue point-range values on the charts, indicate loadings in the same direction, whereas red-colored brain regions and point-range values signify loadings in the opposite direction. Brain images are displayed with NILEARN.

The second LV from this first analysis was associated with lower entorhinal gray matter volume in younger individuals with higher scores on CCI questions 3, 7, 9, and 11, suggesting that these questions (focusing on remembering the recent past, object location, names of family members and friends, and insight into how peers might assess their memory), might be particularly useful for detecting *early* decline associated with entorhinal cortex vulnerability.

### 3.2 Subgroup Analyses Examining Sex and Levels of Education

Two separate PLS analyses were fit to examine how the initial model was impacted by having a high or low education, determined by median split, or being male or female (see Figure 2). Each PLS model yielded a single significant LV, *p*’s < 0.001, and explained 86% and 76% of the cross-block covariance for education group and sex, respectively.

**Figure 2:**
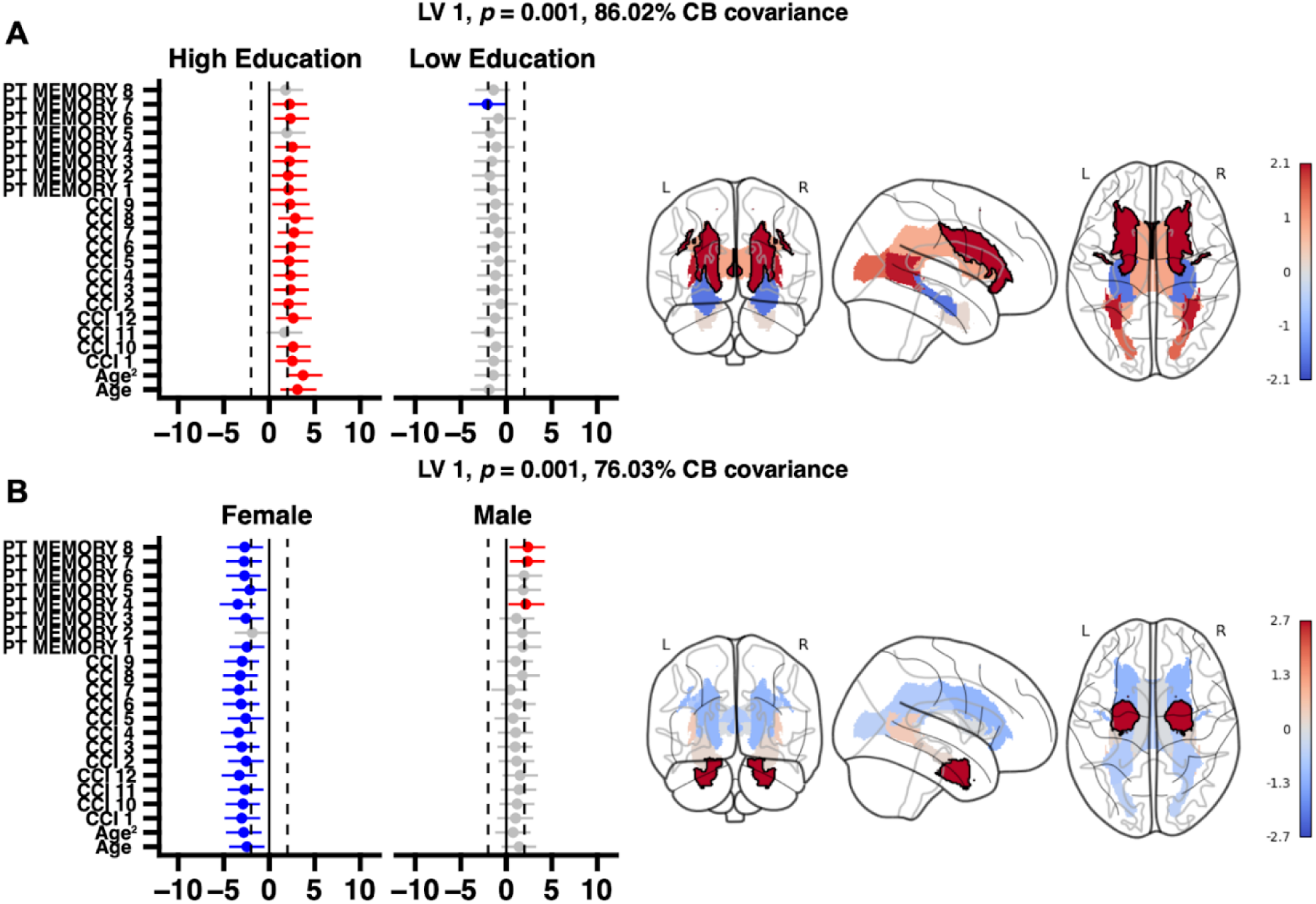
PLS analyses grouped by education and sex. *Notes:* PT_Memory 1 through 8 are the ECOG questions. CCI1 through 12 are the CCI questions. PLS analyses again comparing subjective cognitive decline with brain change; however, this time, the data were grouped by education (A) or sex (B). High versus low education level was determined by median split. Individuals with higher levels of education reliably expressed more white matter hyperintensities in the frontal regions than individuals with lower education, and this was part of a larger (non-reliable) pattern where more WMH and lower hippocampal volumes tended to be associated with greater SCD in the high education group. In Panel B, females who expressed higher levels of subjective cognitive decline and who were older expressed lower entorhinal cortex volumes.

In the education model (see Figure 2A), individuals who are older, and report higher levels of subjective cognitive decline on both the ECog and the CCI have a greater probability of white matter hyperintensities affecting the frontal lobes. A pattern of higher temporal lobe WMH burden and lower hippocampal volume was also associated with this LV, though the bootstrap ratio values for these regions were not reliably different from zero. Thus, more highly educated individuals may have greater insight into their own neural decline than individuals with less education.

In the sex model (Figure 2B), females who were older and had higher values of subjective cognitive decline, measured by either the ECog or the CCI, tended to have reduced volume in the entorhinal cortex. These findings indicate that report of SCD in females may be more strongly associated with structural brain changes than reports of SCD in males.

### 3.3 Brain-Behaviour Correlations

To relate the representativeness of the PLS LVs to behavior, we first extracted latent scores for each individual for the brain and behavioral matrices (i.e., brain and behavioral scores). These scores represent how strongly each individual loaded on the dimension being considered. Next, we used PCA to reduce the dimensionality of six measures of memory (MMSE, RAVLT forgetting, RAVLT learning, RAVLT immediate, MoCA, and ADAS13) to derive a “memory component” (see Figure 3). The first dimension of the PCA explained 39% of the covariance and was primarily driven by the MOCA, ADAS13, and RAVLT immediate scores.

**Figure 3:**
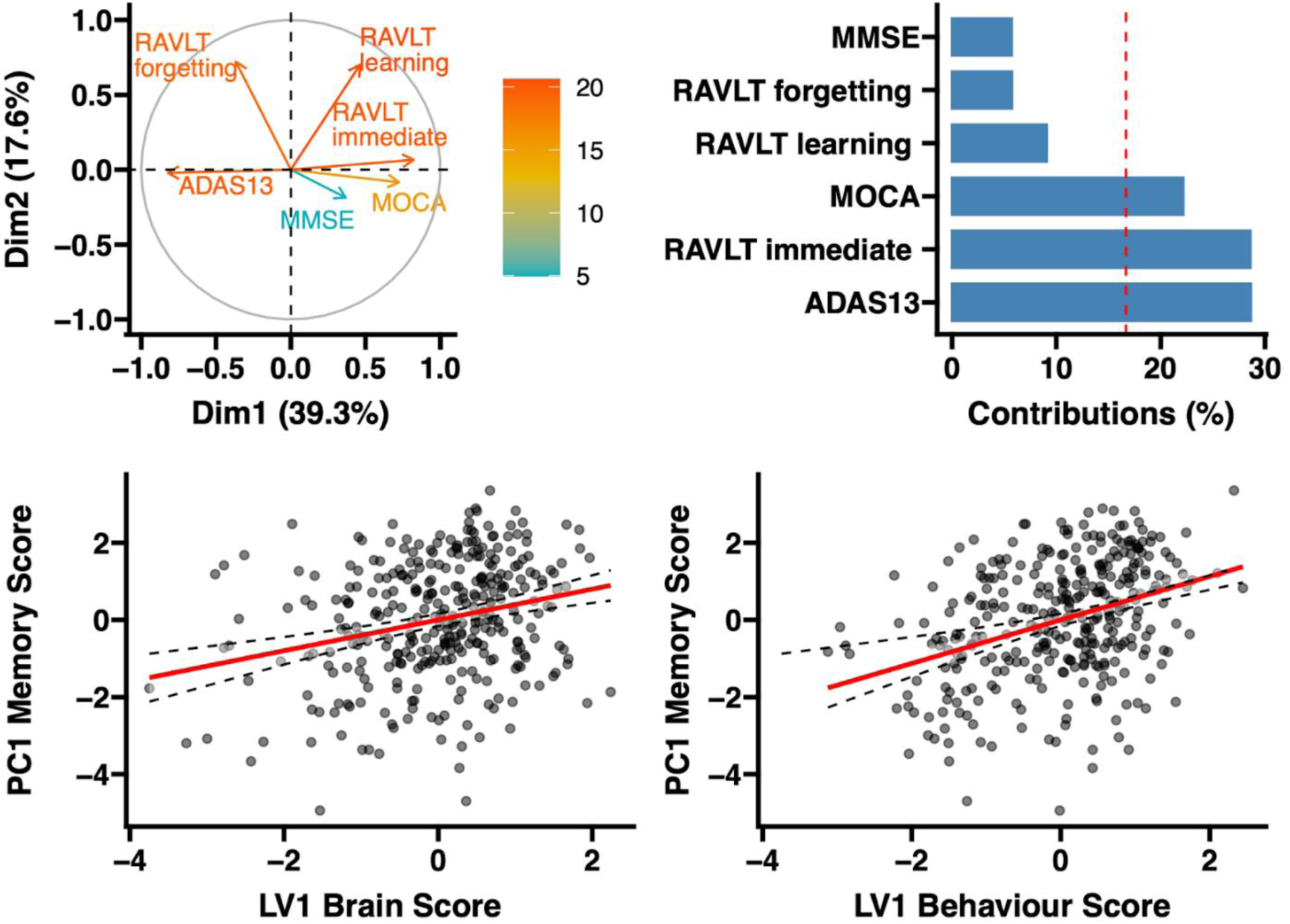
Brain-behaviour correlations with the first latent variable. *Notes*: The brain-behavior correlations observed between the first latent variable (LV1) scores from Partial Least Squares (PLS) analysis of both brain and behavior data, and a composite memory measure. This composite memory measure is derived from a Principal Component Analysis (PCA) of various cognitive assessments, including the Mini-Mental State Examination (MMSE), Rey Auditory Verbal Learning Test (RAVLT) focusing on Forgetting, Learning, and Immediate recall scores, the Montreal Cognitive Assessment (MOCA), and the Alzheimer’s Disease Assessment Scale-Cognitive Subscale-13 (ADAS-13). The first dimension in this analysis captured 39% of the covariance between these variables. Notably, positive scores on this dimension are indicative of better cognitive performance.

We predicted memory performance (using the first component score) with two PLS models using the brain and behaviour scores from the first PLS analysis (see Figure 3). The first model, predicting memory performance from brain structural scores, was significant but only accounted for 7% of the variance in memory performance (R² = 0.07, *F*(1, 332) = 24.02, *p* < .001, adjusted R² = 0.06). The positive and significant effect of brain score on memory performance was quantified with a *β* coefficient of 0.40 (95% CI [0.24, 0.56], *t*(332) = 4.90, *p* < .001), indicating that as brain score increases (i.e., a person expresses a lower probability of white matter hyperintensities and greater hippocampal and entorhinal cortex volume), so does memory performance, albeit to a modest extent.

The second model shifted focus towards behavioral scores, presenting a more robust relationship with the memory component. This model accounted for a moderate 13% of the variance (R² = 0.13, *F*(1, 332) = 51.68, *p* < .001, adjusted R² = 0.13). Similar to the first model, the influence of the behavioural score was both positive and statistically significant (*β* = 0.56, 95% CI [0.41, 0.72], t(332) = 7.19, *p* < .001), suggesting that younger individuals with fewer subjective memory complaints had stronger memory scores.

### 3.4 ROC Analysis

Eighty-seven and 230 individuals had completed one and two year clinical follow up assessments, respectively. Out of those, 15 (17.24%) and 45 (19.6%) individuals experienced cognitive decline in year one and two follow up visits. Figure 4 shows the performance of the first two brain and behaviour LVs as well as the memory PCs in predicting future cognitive decline at years one and two, respectively. The first memory PC was the best differentiator of future cognitive decline, both at year 1 (AUC = 0.80) and year 2 (AUC = 0.75). Brain and behaviour LV scores were also able to differentiate the individuals that demonstrated cognitive decline in follow visits from those that remained stable, albeit to a lesser extent. The best differentiations were observed for LV2 brain score and LV1 behaviour scores at year 1 (AUC = 0.63) and LV1 and LV2 brain scores at year 2 (AUCs = 0.62 and 0.66). Overall, brain scores were better predictors of future cognitive decline (all AUCs > 0.61). Similar results were obtained for a cognitive decline threshold of 1 point increase in CDR (Year 1: N decline= 5; AUC PC1-memory=0.92; AUC LV1-brain=0.78; AUC LV2-brain=0.62, and Year 2: N decline= 22; AUC PC1-memory=0.82; AUC LV1-brain=0.64; AUC LV2-brain=0.65).

**Figure 4.**
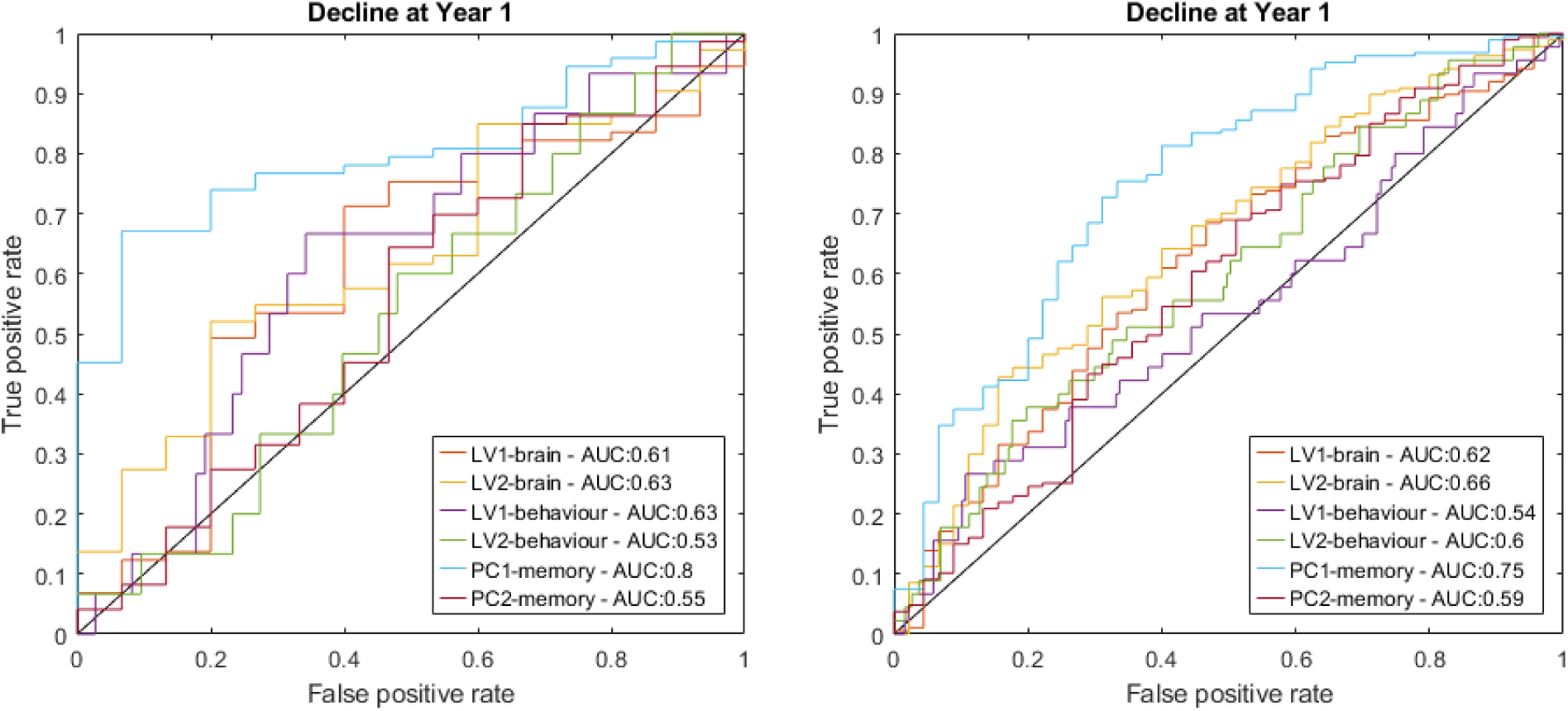
ROC analysis Note: Results are comparing the AUC of PLS LVs in predicting future cognitive decline at one (left panel) and two (right panel) year follow up timepoints.

### 3.5 Secondary Exploratory Analysis

A secondary analysis was also performed with the data regression out age before completing the PLS analysis. This analysis was completed to determine the relationship independent of age. However, it should be noted that both models are important to consider because SCD is strongly associated with age.

In this secondary analysis, one LV was significant, *p* < 0.01 explained 24% of the cross-block covariance. For this LV, 8/12 (67%) CCI variables, but none of the ECoG scores, negatively covaried with entorhinal cortex volume. This finding suggests that above and beyond the effect of age, the CCI is sensitive to early abnormal changes in a region highly sensitive to changes observed in early AD.

Group analyses by education revealed two significant LVs, *p*’s < 0.01, which explained 39% and 24% of the cross-block covariance, respectively. The first LV showed that people with higher education tended to have stronger CCI responses (8/12 CCI scores loaded positively and reliably for this group), than people with less education, (1/12 CCI scores loaded positively for this group). As above, positive scores indicating higher subjective memory complaints were associated with reduced entorhinal cortex volume. The second significant LV was associated with reliably higher frontal and temporal WMH load (BSR values > 2), with generally higher CCI scores in the more educated groups, though these values did not exceed +2, indicating they were less reliable.

After removing the effect of age, re-running the analysis grouped by sex revealed two significant, *p*s < 0.01, LVs that accounted for 65% and 41% of the cross-block covariance, respectively. The first LV revealed that males who had higher CCI scores on questions 1 and 2 and higher ECoG scores on questions 4, 7, and 8 were more likely to have higher WMH burden in frontal, parietal, and occipital lobes, along with larger ventricular volumes. The second LV revealed that CCI scores were reliably associated with entorhinal cortex volume loss in both males (67% of questions) and females (50% of questions).

## 4 Discussion

Much research has determined that SCD is associated with increased rates of cognitive decline (Hohman et al., 2011; Kamberis et al., 2021; Morrison and Oliver, 2022; Rabin et al., 2017) and increases risk for development of AD (Jessen et al., 2020, 2014; Mitchell et al., 2014). However, diverse methodologies exist for classifying SCD, ranging from a single question to extensive questionnaires regarding subjective reports of cognitive change across multiple domains. We previously observed that SCD as endorsed by either the CCI or ECog were associated with different patterns of WMH burden (Morrison et al., 2023) and gray matter volumes(Morrison et al., 2022). To expand on these findings, we examined if the individual questions from these questionnaires mapped onto WMH and gray matter volume to the same degree or if one of the questionnaires were more sensitive to brain changes that are associated with cognitive decline and dementia. Two significant LVs (*p*’s<0.001) explained 85.89% and 8.30% of the cross-block covariance. The first LV identified that older age and higher scores on five questions from the CCI, questions 2,6,7,10,12, were strongly associated with high WMH burden in the frontal, parietal, and temporal region and reductions in gray matter in the hippocampus, entorhinal cortex, and larger ventricles. The second LV shows that younger individuals and those with higher SCD scores on four questions (CCI questions 3,7,9,11) were associated with lower WMH burden in the temporal and parietal lobes and lower entorhinal cortex volumes. To the extent that individuals expressed the brain and behavioral patterns associated with the first latent variable, (i.e., were older, expressed more concern about their memory, and had higher WMH burden and lower hippocampal and entorhinal cortex volume), they also had lower scores on a principal component of memory scores. These results were further explored in males vs. females and in those with higher vs lower education. We observed that almost all questions from both the CCI and ECog (except for question 2 from the ECog) were associated with lower gray matter volume in the entorhinal cortex in females. On the other hand, the questions were not related to reductions in gray matter volume or WMH burden in males. Similarly, individuals with high education expressed more WMH burden in frontal regions compared to low education. These education findings were associated with almost all questions from both the CCI and ECog except ECog questions 5 and 8 and CCI question 11.

The current results show that the CCI questions are sensitive to both volume changes and WMH burden whereas the ECog is not associated with these changes. Although endorsement of memory questions on these two questionnaires have shown a high correlation (*r* =0.75, Wells et al., 2022), our findings suggest that they are not associated with changes in brain structure to the same degree. The results of the current study captured specific associations between CCI questions and neuroimaging markers that are not evident in the ECog questionnaire. These findings are consistent with previous research which observed that there are only moderate correlations between different SCD questionnaires (van Harten et al., 2018; Vogel et al., 2016), which have also concluded that the questionnaires should not be used interchangeably (Vogel et al., 2016). Even within the CCI, our observed findings suggest that only certain questions are indicative or sensitive to the brain changes measured here. It should be noted that when age was removed from the analyses, the main region that remained significantly associated with SCD endorsement was the entorhinal cortex and only with CCI questions (i.e., no ECog questions were significant). This finding suggests that SCD is associated with early AD because the entorhinal cortex is the first brain region to show abnormal changes early in disease progression(Igarashi, 2023). Taken together, the results indicate that the different questionnaires are measuring different constructs and potentially different types of cognitive decline or dementia. Consistent with previous research, further work is needed to develop a harmonized SCD measurement (Butterbrod et al., 2023). From a clinical perspective, excluding questions not linked to age-related brain changes or dementia can streamline appointments, enhancing patient-doctor interaction and efficiency.

When examining if the questions performed differently in males vs. females, we observed that the questions from both CCI and ECog were mainly associated with lower gray matter volume in females. Given that entorhinal cortex is consistently reported as one of the first brain regions to show abnormal changes in AD (Igarashi, 2023), these findings suggest that female with SCD may be on the trajectory for AD-related brain changes. This finding is consistent with previous reports showing that SCD is more strongly associated with cognitive decline in females than males over 15 years (Oliver et al., 2022) and that females with SCD are more likely to convert to dementia (Heser et al., 2019). Even when females exhibit the same amount of age- and dementia-related brain changes as males they experience more cognitive decline and worse dementia outcomes (Barnes et al., 2005; Ferretti et al., 2018). Therefore, future research and interventions should focus on interventions for females who report SCD to help reduce cognitive decline, increased WMH burden, and gray matter reductions.

In addition to variable results in males vs. females, differences in the questions sensitivity to brain changes were also observed in those with high vs. low education. The findings observed here suggest that those with higher education who endorse questions on both the CCI and ECog indicating SCD, exhibit more WMH burden in frontal regions and this was part of a larger (non-reliable) pattern where more WMH and lower hippocampal volumes tended to be associated with greater SCD in the high education group. On the other hand, endorsement of the questions in people with lower education was not sensitive to brain changes. Older adults with higher education may thus be more sensitive to early changes in cognitive function than those with lower education. As a result of CR, higher-educated older adults require more brain changes before clinical symptoms are apparent than those with lower education. People with SCD who are highly educated may thus already have a large amount of structural brain changes before they start experiencing subtle changes in cognition, therefore once they endorse SCD they already exhibit more brain changes than someone with lower education. This interpretation is consistent with previous work examining other pathological brain changes in people with SCD, which observed that the relationship between SCD and amyloid burden is stronger in those with higher education (Aghjayan et al., 2017) and that those with higher education and SCD had a greater risk of decline to AD than those with lower education (van Oijen et al., 2007).

The current study also revealed a significant association between brain and behaviour. Firstly, we observed that high brain scores, indicating lower probability of WMHs and greater hippocampal and entorhinal cortex volume, were associated with better memory performance, albeit to a modest extent. These findings are consistent with previous work showing neurodegeneration in the medial temporal lobe are associated with impairments in memory functioning (for review see, Fjell et al., 2014)). Importantly, these brain-behaviour analyses also revealed that younger individuals with fewer subjective memory complaints (i.e., less SCD) had stronger memory scores. Finally, the ROC curve analyses suggested that PLS brain and behaviour scores are predictive of future cognitive decline in individuals with SCD.

Overall, our findings suggest that overall SCD endorsed by CCI is more sensitive to WMH burden and hippocampal, entorhinal cortex, and ventricle volume than SCD endorsed by the ECog. However, these patterns differ based on sex and level of education, with both the questionnaires being more sensitive to entorhinal cortex volume loss in females than males and to WMH burden in the frontal region in those with high education compared to those with low education. These findings underscore the significance of differentiating between subjective cognitive decline endorsed by different assessment tools. Understanding these nuances is crucial for both clinical practice and research endeavors. Further investigations are warranted to elucidate the specific questionnaire items associated with dementia subtypes, facilitating early prediction and treatment options.

## Data Availability

All data produced are available online at adni.loni.usc.edu

https://adni.loni.usc.edu/

## Notes

**Funding:** CM supported by the Canadian Institutes of Health Research (CIHR), JAEA is supported by CRC II (CRC-2020-00174), NSERC-DG (DGECR-2022-00309), MD is supported by the Healthy Brains for Healthy Lives (HBHL), Alzheimer Society Research Program (ASRP), Natural Sciences and Engineering Research Council of Canada (NSERC), Fonds de Recherche du Québec (FRQS), Douglas Research Centre (DRC) and CIHR.

**Disclosure statement:** The authors have no conflicts of interest to disclose.

### Competing Interest Statement

The authors have declared no competing interest.

### Funding Statement

CM supported by the Canadian Institutes of Health Research (CIHR), JAEA is supported by CRC II (CRC-2020-00174), NSERC-DG (DGECR-2022-00309), MD is supported by the Healthy Brains for Healthy Lives (HBHL), Alzheimer Society Research Program (ASRP), Natural Sciences and Engineering Research Council of Canada (NSERC), Fonds de Recherche du Quebec (FRQS), Douglas Research Centre (DRC) and CIHR.

### Author Declarations

The study used data that are openly available at teh adni.loni.usc.edu website.

## References

1. Aghjayan, S.L., Buckley, R.F., Vannini, P., Rentz, D.M., Jackson, J.D., Sperling, R.A., Johnson, K.A., Rebecca, E., Hospital, M.G., Hospital, M.G., Medicine, N., Hospital, G., 2017. The influence of demographic factors on subjective cognitive concerns and beta-amyloid 29, 645–652. 10.1017/S1041610216001502.The

2. Anderson, J.A.E., Hawrylewicz, K., Grundy, J.G., 2020. Does bilingualism protect against dementia? A meta-analysis. Psychonomic Bulletin and Review 27, 952–965. 10.3758/s13423-020-01736-5

3. Anor, C.J., Dadar, M., Collins, D.L., Tartaglia, M.C., 2021. The Longitudinal Assessment of Neuropsychiatric Symptoms in Mild Cognitive Impairment and Alzheimer’s Disease and Their Association With White Matter Hyperintensities in the National Alzheimer’s Coordinating Center’s Uniform Data Set. Biological Psychiatry: Cognitive Neuroscience and Neuroimaging 6, 70–78. 10.1016/j.bpsc.2020.03.006

4. Barnes, L.L., Wilson, R.S., Bienias, J.L., Schneider, J.A., Evans, D.A., Bennett, D.A., 2005. Sex differences in the clinical manifestations of Alzheimer disease pathology. Archives of General Psychiatry 62, 685–691. 10.1001/archpsyc.62.6.685

5. Beaton, D., Chin Fatt, C.R., Abdi, H., 2014. An ExPosition of multivariate analysis with the singular value decomposition in R. Computational Statistics and Data Analysis 72, 176–189. 10.1016/j.csda.2013.11.006

6. Butterbrod, E., Rabin, L., Tommet, D., Jones, R.N., Dubbelman, M.A., Crane, P.K., Sikkes, S.A.M., 2023. Toward optimizing measurement of self-perceived cognitive function - a global expert survey on behalf of the SCD-I Working Group. Alzheimer’s & Dementia.

7. Coupe, P., Yger, P., Prima, S., Hellier, P., Kervrann, C., Barillot, C., 2008. An optimized blockwise nonlocal means denoising filter for 3-D magnetic resonance images. IEEE Transactions on Medical Imaging 27, 425–441. 10.1109/TMI.2007.906087

8. Crook, T.H., Feher, E.P., Larrabee, G.J., 1992. Assessment of Memory Complaint in Age-Associated Memory Impairment: The MAC-Q. International Psychogeriatrics 4, 165–176. 10.1017/S1041610292000991

9. Dadar, M., Fereshtehnejad, S.M., Zeighami, Y., Dagher, A., Postuma, R.B., Collins, D.L., 2020. White Matter Hyperintensities Mediate Impact of Dysautonomia on Cognition in Parkinson’s Disease. Movement Disorders Clinical Practice 7, 639–647. 10.1002/mdc3.13003

10. Dadar, M., Fonov, V.S., Collins, D.L., 2018. A comparison of publicly available linear MRI stereotaxic registration techniques. NeuroImage 174, 191–200. 10.1016/j.neuroimage.2018.03.025

11. Dadar, M., Maranzano, J., Ducharme, S., Collins, D.L., 2019. White matter in different regions evolves differently during progression to dementia. Neurobiology of Aging 76, 71–79. 10.1016/j.neurobiolaging.2018.12.004

12. Dadar, M., Misquitta, K., Anor, C.J., Fonov, V.S., Tartaglia, M.C., Carmichael, O.T., Decarli, C., Collins, D.L., 2017a. NeuroImage Performance comparison of 10 di ff erent classi fi cation techniques in segmenting white matter hyperintensities in aging 157, 233–249. 10.1016/j.neuroimage.2017.06.009

13. Dadar, M., Pascoal, T.A., Manitsirikul, S., Misquitta, K., Fonov, V.S., Carmela, M., Breitner, J., Rosa-neto, P., Carmichael, O.T., Decarli, C., Collins, D.L., 2017b. Validation of a Regression Technique for Segmentation of White Matter Hyperintensities in Alzheimer’s Disease 36, 1758–1768.

14. Farias, S.T., Mungas, D., Reed, B.R., Cahn-Weiner, D., Jagust, W., Baynes, K., DeCarli, C., 2008. The Measurement of Everyday Cognition (ECog): Scale Development and Psychometric Properties. Neuropsychology 22, 531–544. 10.1037/0894-4105.22.4.531

15. Ferretti, M.T., Iulita, M.F., Cavedo, E., Chiesa, P.A., Dimech, A.S., Chadha, A.S., Baracchi, F., Girouard, H., Misoch, S., Giacobini, E., Depypere, H., Hampel, H., 2018. Sex differences in Alzheimer disease — The gateway to precision medicine. Nature Reviews Neurology 14, 457–469. 10.1038/s41582-018-0032-9

16. Fjell, A.M., McEvoy, L., Holland, D., Dale, A.M., Walhovd, K.B., 2014. What is normal in normal aging? Effects of aging, amyloid and Alzheimer’s disease on the cerebral cortex and the hippocampus. Progress in Neurobiology 117, 20–40. 10.1016/j.pneurobio.2014.02.004

17. Fonov, V., Evans, A.C., Botteron, K., Almli, C.R., McKinstry, R.C., Collins, D.L., 2011. Unbiased average age-appropriate atlases for pediatric studies. NeuroImage 54, 313–327. 10.1016/j.neuroimage.2010.07.033

18. Gauthier, S., Albert, M., Fox, N., Goedert, M., Kivipelto, M., Mestre-Ferrandiz, J., Middleton, L.T., 2016. Why has therapy development for dementia failed in the last two decades? Alzheimer’s and Dementia 12, 60–64. 10.1016/j.jalz.2015.12.003

19. Hammers, A., Allom, R., Koeep, M., Free, S., Myers, R., Lemieux, L., Mitchell, T., Brooks, D., Duncan, J., 2003. Validation of T1w-based segmentations of white matter hyperintensity volumes in large-scale datasets of aging. Human brain mapping 19, 224–247.

20. Heser, K., Kleineidam, L., Wiese, B., Oey, A., Roehr, S., Pabst, A., Kaduszkiewicz, H., van den Bussche, H., Brettschneider, C., König, H.-H., Weyerer, S., Werle, J., Fuchs, A., Pentzek, M., Mösch, E., Bickel, H., Maier, W., Scherer, M., Riedel-Heller, S., Wagner, M., 2019. Subjective Cognitive Decline May Be a Stronger Predictor of Incident Dementia in Women than in Men. Jouirnal of Alzheimer’s Disease 68, 1469–1478.

21. Hohman, T.J., Beason-Held, L.L., Lamar, M., Resnick, S.M., 2011. Subjective Cognitive Complaints and Longitudinal Changes in Memory and Brain Function Timothy. Neuropsychology 25, 125–130. 10.1037/a0020859.Subjective

22. Igarashi, K.M., 2023. Entorhinal cortex dysfunction in Alzheimer’s disease. Trends in Neurosciences 46, 124–136. 10.1016/j.tins.2022.11.006

23. Jessen, F., Amariglio, R., van Boxtel, M.P.J., Breteler, M.M., Ceccaldi, M., Chetelat, G., Dubois, B., Dufouil, C., 2014. A conceptual framework for research on subjective cognitive decline in preclinical Alzheimer’s disease 10, 844–852. 10.1016/j.jalz.2014.01.001.A

24. Jessen, F., Amariglio, R.E., Buckley, R.F., Flier, W.M.V.D., Han, Y., Molinuevo, J.L., Rabin, L., Rentz, D.M., 2020. The characterisation of subjective cognitive decline 271–278.

25. Jessen, F., Feyen, L., Freymann, K., Tepest, R., Maier, W., Heun, R., Schild, H.H., Scheef, L., 2006. Volume reduction of the entorhinal cortex in subjective memory impairment. Neurobiology of Aging 27, 1751–1756. 10.1016/j.neurobiolaging.2005.10.010

26. Josse, J., Husson, F., 2016. missMDA: A package for handling missing values in multivariate data analysis. Journal of Statistical Software 70. 10.18637/jss.v070.i01

27. Kamberis, N., Cavuoto, M.G., Pike, K.E., 2021. The influence of subjective cognitive decline on prospective memory over 5 years. Neuropsychology 35, 78–89. 10.1037/neu0000709

28. Krishnan, A., Williams, L.J., McIntosh, A.R., Abdi, H., 2011. Partial Least Squares (PLS) methods for neuroimaging: A tutorial and review. NeuroImage 56, 455–475. 10.1016/j.neuroimage.2010.07.034

29. McIntosh, A.R., Lobaugh, N.J., 2004. Partial least squares analysis of neuroimaging data: Applications and advances. NeuroImage 23, 250–263. 10.1016/j.neuroimage.2004.07.020

30. Mitchell, A.J., Beaumont, H., Ferguson, D., Yadegarfar, M., Stubbs, B., 2014. Risk of dementia and mild cognitive impairment in older people with subjective memory complaints: metaanalysis. Acta psychiatrica Scandinavica 130, 439–451.

31. Morrison, C., Dadar, M., Shafiee, N., Villeneuve, S., Collins, D.L., 2022. Regional brain atrophy and cognitive decline depend on definition of subjective cognitive decline. NeuroImage: Clinical 33. 10.1016/j.nicl.2021.102923

32. Morrison, C., Dadar, M., Villeneuve, S., Ducharme, S., Collins, D.L., 2023. White matter hyperintensity load varies depending on subjective cognitive decline criteria. GeroScience 45, 17–28. 10.1007/s11357-022-00684-3

33. Morrison, C., Oliver, M., 2022. Subjective Cognitive Decline Is Associated With Lower Baseline Cognition and Increased Rate of Cognitive Decline. The Journals of Gerontology: Series B.

34. Morrison, C., Dadar, M., Collins, D.L., 2023. Sex differences in risk factors, burden, and outcomes of cerebrovascular disease in Alzheimer’s disease populations. Alzheimer’s & Dementia.

35. Oliver, M.D., Morrison, C., Kamal, F., Graham, J., Dadar, M., 2022. Subjective cognitive decline is a better marker for future cognitive decline in females than in males. Alzheimer’s Research and Therapy 14, 1–12. 10.1186/s13195-022-01138-w

36. Perrotin, A., La Joie, R., de La Sayette, V., Barré, L., Mézenge, F., Mutlu, J., Guilloteau, D., Egret, S., Eustache, F., Chételat, G., 2017. Subjective cognitive decline in cognitively normal elders from the community or from a memory clinic: Differential affective and imaging correlates. Alzheimer’s and Dementia 13, 550–560. 10.1016/j.jalz.2016.08.011

37. Rabin, L.A., Smart, C.M., Amariglio, R.E., 2017. Subjective Cognitive Decline in Preclinical Alzheimer’s Disease. Annual Review of Clinical Psychology 13, 369–396. 10.1146/annurev-clinpsy-032816-045136

38. Reisberg, B., Shulman, M.B., Torossian, C., Leng, L., Zhu, W., 2010. Outcome over seven years of healthy adults with and without subjective cognitive impairment 6, 11–24. 10.1016/j.jalz.2009.10.002

39. Rooden, S.V., Berg-huysmans, A.A.V.D., Croll, P.H., Labadie, G., 2018. Subjective Cognitive Decline Is Associated with Greater White Matter Hyperintensity Volume 66, 1283–1294. 10.3233/JAD-180285

40. Saykin, A.J., Wishart, H.A., Rabin, L.A., Santulli, R.B., Flashman, L.A., West, J.D., McHugh, T.L., Mamourian, A.C., 2006. Older adults with cognitive complaints show brain atrophy similar to that of amnestic MCI. Neurology 67, 834–842. 10.1212/01.wnl.0000234032.77541.a2

41. Sled, J.G., Zijdenbos, A.P., Evans, A.C., 1998. A nonparametric method for automatic correction of intensity nonuniformity in mri data. IEEE Transactions on Medical Imaging 17, 87–97. 10.1109/42.668698

42. Snitz, B.E., Lopez, O.L., McDade, E., Becker, J.T., Cohen, A.D., Price, J.C., Mathis, C.A., Klunk, W.E., 2015. Amyloid-beta imaging in older adults presenting to a memory clinic with subjective cognitive decline: A pilot study. Journal of Alzheimer’s Disease 48, S151–S159. 10.3233/JAD-150113

43. Sperling, R.A., Aisen, P.S., 2011. Testing the Right Target and Right Drug at the Right Stage 3, 1–5.

44. Stern, Y., 2013. Cognitive reserve in ageing. Lancet Neurol. 11, 1006–1012. 10.1016/S1474-4422(12)70191-6.Cognitive

45. Stern, Y., 2002. What is cognitive reserve? Theory and research application of the reserve concept. Journal of the International Neuropsychological Society 8, 448–460. 10.1017/S1355617702813248

46. Striepens, N., Scheef, L., Wind, A., Popp, J., Spottke, A., Cooper-Mahkorn, D., Suliman, H., Wagner, M., Schild, H.H., Jessen, F., 2010. Volume loss of the medial temporal lobe structures in subjective memory impairment. Dementia and Geriatric Cognitive Disorders 29, 75–81. 10.1159/000264630

47. van Harten, A.C., Mielke, M.M., Swenson-dravis, D.M., Hagen, C.E., 2018. Subjective cognitive decline and risk of MCI The Mayo Clinic Study of Aging. 10.1212/WNL.0000000000005863

48. van Oijen, M., de Jong, F.J., Hofman, A., Koudstaal, P.J., Breteler, M.M.B., 2007. Subjective memory complaints, education, and risk of Alzheimer’s disease. Alzheimer’s and Dementia 3, 92–97. 10.1016/j.jalz.2007.01.011

49. Vogel, A., Salem, L.C., Andersen, B.B., Waldemar, G., 2016. Differences in quantitative methods for measuring subjective cognitive decline - Results from a prospective memory clinic study. International Psychogeriatrics 28, 1513–1520. 10.1017/S1041610216000272

50. Wang, X., Huang, W., Su, L., Xing, Y., Jessen, F., Sun, Y., Shu, N., 2020. Neuroimaging advances regarding subjective cognitive decline in preclinical Alzheimer’s disease 3, 1–27.

51. Wells, L.F., Risacher, S.L., McDonald, B.C., Farlow, M.R., Brosch, J., Gao, S., Apostolova, L.G., Saykin, A.J., 2022. Measuring Subjective Cognitive Decline in Older Adults: Harmonization between the Cognitive Change Index and the Measurement of Everyday Cognition Instruments. Journal of Alzheimer’s Disease 87, 761–769. 10.3233/JAD-215388

52. Wilson, R.S., Boyle, P.A., Yang, J., James, B.D., Bennett, D.A., 2015. Early Life Instruction in Foreign Language and Music and Incidence of Mild Cognitive Impairment. Neuropsychology 29, 292–302.

